# Factors associated with human papillomavirus (HPV) virus load variations in genital infections in young women

**DOI:** 10.1101/2024.01.17.24301437

**Authors:** Nicolas Tessandier, Vanina Boué, Tsukushi Kamyia, Olivier Supplisson, Carmen Lia Murall, Bastien Reyné, Christian Selinger, Claire Bernat, Sophie Grasset, Soraya Groc, Massilva Rahmoun, Marine Bonneau, Vincent Foulongne, Christelle Graf, Vincent Tribout, Jean-Luc Prétet, Jacques Reynes, Michel Segondy, Ignacio G Bravo, Nathalie Boulle, Samuel Alizon

## Abstract

Human papillomaviruses (HPVs) are the most oncogenic viruses known to humans, with 12 high-risk (HR) genotypes causing nearly all cervical cancers. Cytology is commonly used to screen for cervical lesions but is currently being replaced by testing for high-risk HPV (HR HPV). Although HR HPV screening has a higher sensitivity, its specificity is limited, and it is currently advised to repeat the first screening 4 to 6 months later. To increase the sensitivity of the screening triage, other biomarkers have been suggested, including HPV viral load. Indeed, since 1999, several independent studies have found an association between HR HPV viral load in cervical samples and the severity of cervical disease. Here, we further explore the determinants of variations in HPV viral load in genital infections in young adult women.

We analysed samples collected in the PAPCLEAR clinical cohort for participants who were infected by HPV genotypes for which we quantified virus load using qPCR targeting 13 genotypes. We developed a Bayesian statistical model estimating the effect of covariates of interest on the HPV viral load. To analyse precisely the viral load difference between HPV genotypes, phylogenetic distances between HPVs were also integrated in the Bayesian model.

Our results fail to identify an effect of anti-HPV vaccination, co-infections by multiple HPVs or tobacco smoking on the detected viral load. On the opposite, swabs contained significantly more viral copies than cervical smears. Our results also highlight that most of the viral load variance could be explained at the genotype level (80%) rather than at the individual level (20%). Our model reveals important differences in viral load detected between the different genotypes tested, with HPV16 being the highest and HPV18 the lowest. The impact of phylogenetic signal on viral load was also estimated to be low, except for a cluster comprised of HPV53, HPV66 and HPV56. These results contribute to identifying the main drivers of HPV viral load detected and could help design needed future screening policies.

## 1. Introduction

Human papillomaviruses (HPVs) are the most oncogenic viruses known to humans, with 12 high-risk (HR) genotypes causing nearly all cervical cancers and a large fraction of other ano-genital or head-and-neck cancers (de Martel et al. 2017). HPVs are also highly transmissible and represent one of the most prevalent sexually transmitted infections (STIs), affecting more than 80% of sexually active individuals before the age of 45 according to epidemiological models (Chesson et al. 2014).

Cervical cancer prevention relies on cytology screening to identify pre-cancerous lesions and, more recently, on vaccination. The former is still commonly used but is increasingly replaced in first-line screening by high-risk HPV (HR HPV) detection, especially for women older than 30. HR HPV screening has a high sensitivity but a low specificity as a cancer predictor. One of the reasons is that nearly 40% of women positive for HR HPV screening clear the infection within 4 to 6 months, which is why policies tend to implement a second test in this time frame (Gustavsson et al. 2018; Ramanakumar et al. 2016). If the HR HPV remains positive at the second test, a cytological examination by a pathologist is recommended (Saslow et al. 2012).

Devising a single primary screening test able to distinguish transient from persistent infections (or even precancerous lesions) would be particularly useful in the field to reduce the number of tests, improve the screening coverage, and reduce the need for trained specialists. Several biomarkers have been suggested, including methylation of HPV genes (Hillyar et al. 2022) or immunohistochemical staining of the cervical smears (Sun et al. 2019). A third biomarker is HPV viral load in cervical samples, which has been found to be associated with the severity of cervical disease in several independent studies (Hernández-Hernández et al. 2003; Manawapat-Klopfer et al. 2018; Swan et al. 1999; Wu et al. 2017). The risk of cervical cancer or high-grade squamous intraepithelial lesion (HSIL) also increases with the viral load. Another study shows that HR HPV viral load in the primary screening HPV test is associated with the presence of cervical lesions and could be used in triaging HR-HPV positive women for different follow-up strategies or recall times (Berggrund et al. 2019). Another report that viral load is significantly more elevated in patients with HSIL for some specific genotypes (including HPV16) whereas for other genotypes viral load of other genotypes was not (Dong et al. 2018). A recent study conducted on a French cohort found similar findings (Baumann et al. 2021). This is why some have put forward this biomarker to help the triage women testing positive for HR HPV infection (Castle et al. 2002; Sun et al. 2001).

Some of the variations in HPV viral load may be explained by the biology of the infection. For instance, lower levels of HPV16 and HPV18 have been reported in coinfected patients, especially when related HPV genotypes are involved in the co-infection (Xi et al. 2009). Furthermore, interactions between HR and low-risk HPVs could decrease the risk of cervical cancer (Sundström et al. 2015). There is also evidence that vaccination reduces the virus load of non-vaccine genotypes but only for some genotypes and in a limited way (Weele et al. 2019).

Other studies have investigated the link between virus load and the course of the infection. For instance, in a Dutch cohort of women aged 16-29 years, baseline viral load for HPV16 and HPV18 were higher in persistent infections, and the authors hypothesised that HR HPV viral load could be used as a biomarker to distinguish between progression to cervical lesions or regression of the infection (van der Weele et al. 2016). Before that, several studies had proposed to use serial genotype-specific HPV viral load measurement to distinguish regressing cervical lesions from serial virion productive transient infections (Chang et al. 2014; Depuydt et al. 2015, 2016). Another study used 2 to 3 consecutive measurements to predict the outcome of an HR HPV infection, including the grade of the subsequent cervical lesions (Verhelst et al. 2017).

Here, we explore the determinants of variations in HPV viral load in genital infections in young adult women (18 to 25). We analyse qPCR data from a longitudinal cohort of 98 women infected by at least one HPV genotype followed twice at a clinic in 4 weeks, with additional weekly self-samples performed between the visits. Our study stands out by the combination of several factors. First, we have detailed metadata about the participants as well as different types of samples collected during the same visit. Second, we use a sensitive quantitative PCR (qPCR) assay that specifically targets 13 genotypes. Finally, we use elaborate statistical modelling that also accounts for the genomic distance between genotypes. This allows us to show that some sampling methods are associated with higher virus load and to detect significant differences between HPV genotypes, especially HPV16.

## 2. Results

### Study population

We analysed cervical smears, vaginal swabs, and metadata from 98 HPV-positive participants (Table 1). 53 participants (54%) were infected by a single HPV while 45 (46%) were co-infected by multiple HPV genotypes. In line with our earlier results on a subset of this cohort (Murall et al. 2020), co-infected participants were less vaccinated, although the difference was non-significant (35.6 vs 56.6 %, *p* = 0.06), and had a significantly lower body mass index (BMI) than HPV-negative participants (*p* = 0.024). Mono-infected and co-infected had comparable lifetime number of partners and duration of follow-up (Table 1).

**Table 1.** Cohort participants main characteristics stratified by HPV status (uninfected, singly-infected, or co-infected). Differences between groups are tested using a Kruskal-Wallis test.

|  | HPV mono-infection | HPV co-infection | p |
| --- | --- | --- | --- |
| n | 53 | 45 |  |
| Vaccinated against HPV = Yes (n (%)) | 30 (56.6) | 16 (35.6) | 0.060 |
| Lifetime number of partners (mean (SD)) | 12.00 (13.16) | 14.31 (9.76) | 0.333 |
| Duration of follow up (days) (mean (SD)) | 304.26 (225.36) | 327.02 (211.60) | 0.610 |
| Age at first visit (mean (SD)) | 21.64 (2.05) | 21.42 (2.02) | 0.596 |
| Age at menarchy (mean (SD)) | 12.69 (1.48) | 13.13 (1.38) | 0.133 |
| First intercourse (age) (mean (SD)) | 16.13 (1.63) | 16.69 (2.17) | 0.151 |
| **BMI (mean (SD))** | **23.07 (3.66)** | **21.43 (3.38)** | **0.024** |
| Smoking (n (%)) |  |  | 0.473 |
| No | 38 (71.7) | 27 (60.0) |  |
| Occasionally | 4 (7.5) | 5 (11.1) |  |
| Regularly | 11 (20.8) | 13 (28.9) |  |

Among the participants co-infected by several HPV genotypes, the most frequently detected genotypes were HPV51 and HPV53, followed by HPV66 (see Supplementary Tables S2 and S3 for the detailed distribution). The majority of co-infected participants were infected by 2 different genotypes (*n* = 22) and the maximum number of genotypes detected was 6 (*n* = 1) (Figure S3).

To study the impact of co-variates on HPV viral load, we selected a subset based on their potential to be used in clinical diagnostic settings. As detailed in the Methods, we developed a Bayesian hierarchical model with 12 fixed effects and 3 random effects (one at the individual level, another at the genotype level, and a third one based on the HPV phylogenetic distance using the variance:covariance matrix extracted from figure S1).

The type of samples had the strongest impact on HPV viral load, with vaginal swab samples collected in Amies medium having on average a higher estimated HPV viral load compared to cervical smears collected in PBS (1.23 [0.35, 2.11]). Cervical smears collected in Thinprep displayed a tendency toward higher estimated viral load, but to a lower extent (0.51 [−0.13, 1.16]). Vaginal swabs also displayed a lower variance, suggesting a more consistent estimation of the viral load (−0.23 [−0.38, −0.07]). There was also a slight effect of the person performing the sample on the resulting virus load.

On the biological side, the composition of the vaginal microbiota slightly affects the virus load with CST-IV (Lactobacillus-poor) being associated with lower HPV virus loads (−0.53 [−1.29, 0.23]). Increased age or reporting recent menses were also associated with lower virus loads. Other covariates such as smoking status or vaccination status had no detectable impact on estimated HPV viral load.

To explore potential differences in viral load per HPV genotype, we analysed the posterior viral load distributions from the ‘genotype’ random effect in our model (see the Methods). HPV16 was the genotype with the highest estimated posterior viral load, whereas HPV18 was second to last. Overall, the mean difference between the highest and the lowest estimated posterior viral loads (HPV16 and HPV35) spanned across 5 logs. Of note, HPV53 and HPV66, the only two HPV in our dataset belonging to the *2b* IARC risk category, displayed a relatively high posterior viral load, suggesting that the relationship between viral load and carcinogenic risk is not trivial, with the exception of HPV16.

Since the sampling technique was one of the covariates displaying the strongest impact on HPV estimated viral load (Figure 1), we further investigate this issue for all the HPV genotypes for which all three sample types (vaginal swab in Amies, cervical smear in PBS, and cervical smear in Thinprep) were available. Comparisons on a per-genotype basis using the raw viral load (not normalised by albumin) showed that our result that Amies vaginal swabs had higher viral load than cervical smears was largely driven by HPV16. In some other cases, e.g. HPV39, the effect was very limited. In others, such as HPV52, HPV53, or HPV56, Amies vaginal swabs displayed higher viral loads than cervical smears, albeit to a lower extent than for HPV16. Finally, in few cases like HPV66, Amies vaginal swabs collected and PBS cervical smears displayed a higher detected viral load than Thinprep cervical smears. Overall, the genotype seems to have a strong effect on the association between the type of samples and the viral load.

**Figure 1.**
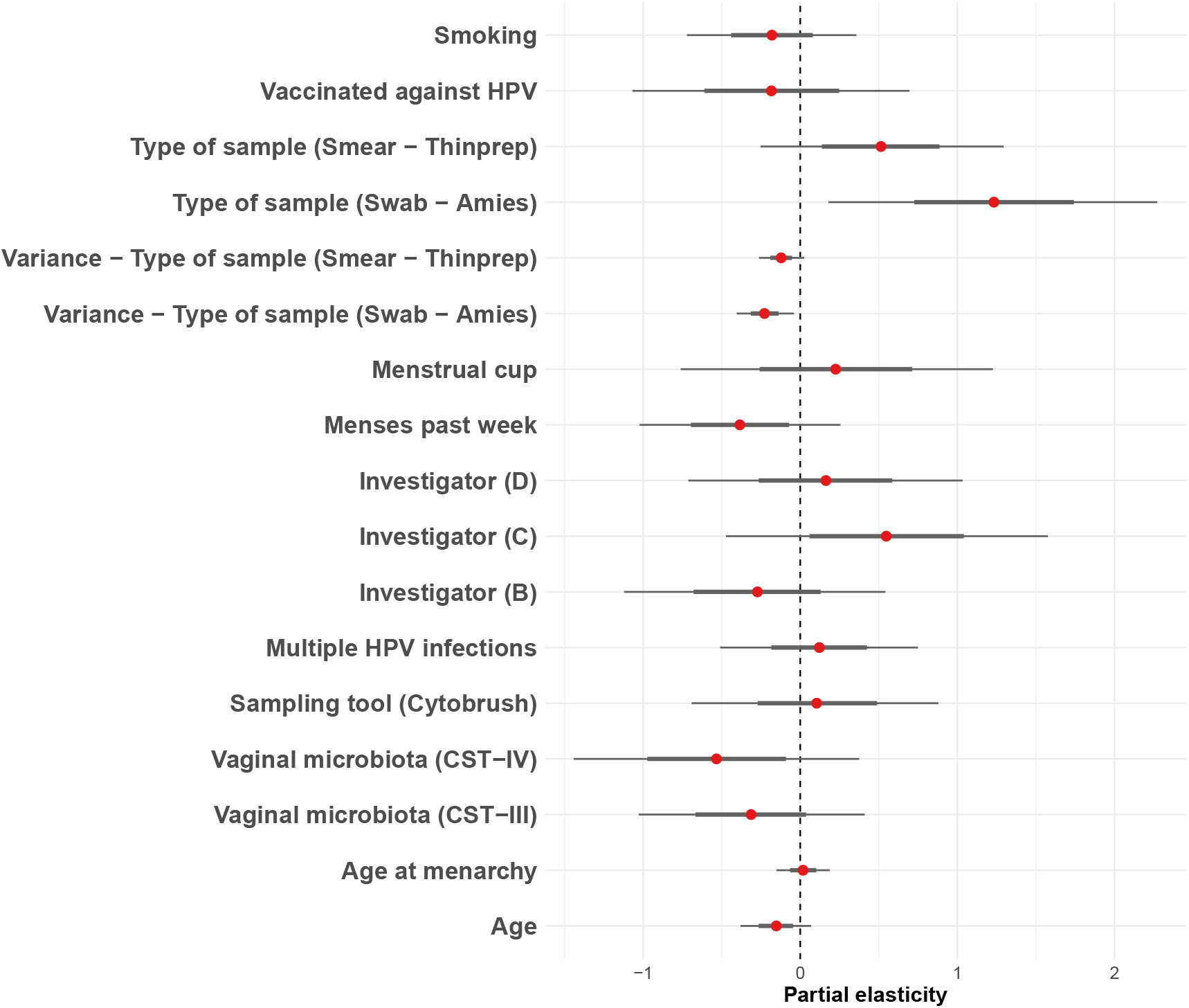
Effect of selected covariates on HPV viral load. We estimated a partial elasticity for each covariate included in the model (see the Methods for details). Red dots indicate posterior medians, thick lines represent 66% quantile intervals (or percentiles), and thin lines 95% quantile intervals.

**Figure 2.**
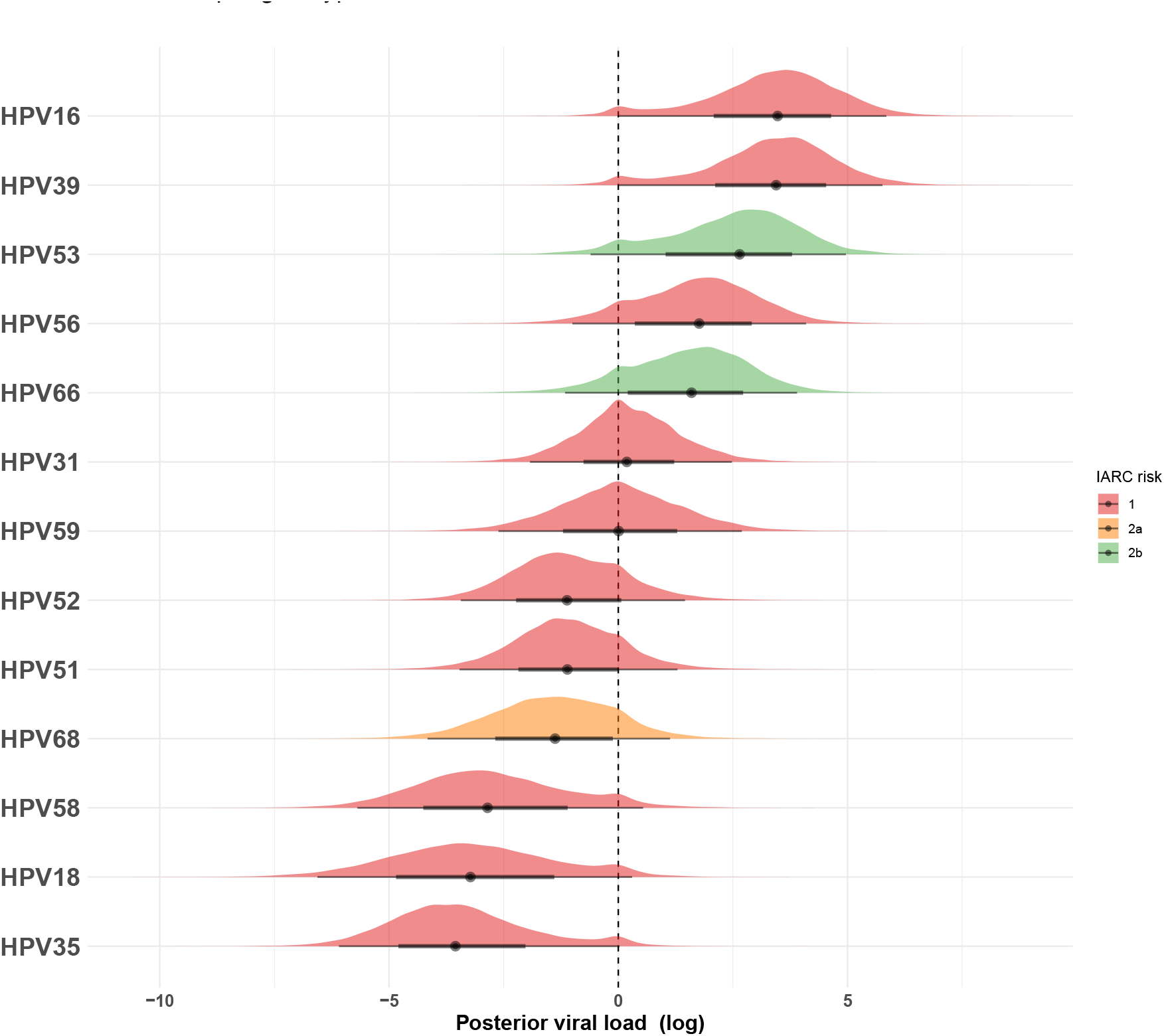
Estimated HPV viral load per genotype. For each genotype included in the model, we estimated a posterior viral load distribution using the cumulative distribution function from the ggdist package. Colors are based on the IARC 2012 oncogeneticity risk IARC Working Group on the Evaluation of Carcinogenic Risks to Humans (2012). Thick and thin lines show the estimated 66% and 95% quantiles intervals respectively.

**Figure 3.**
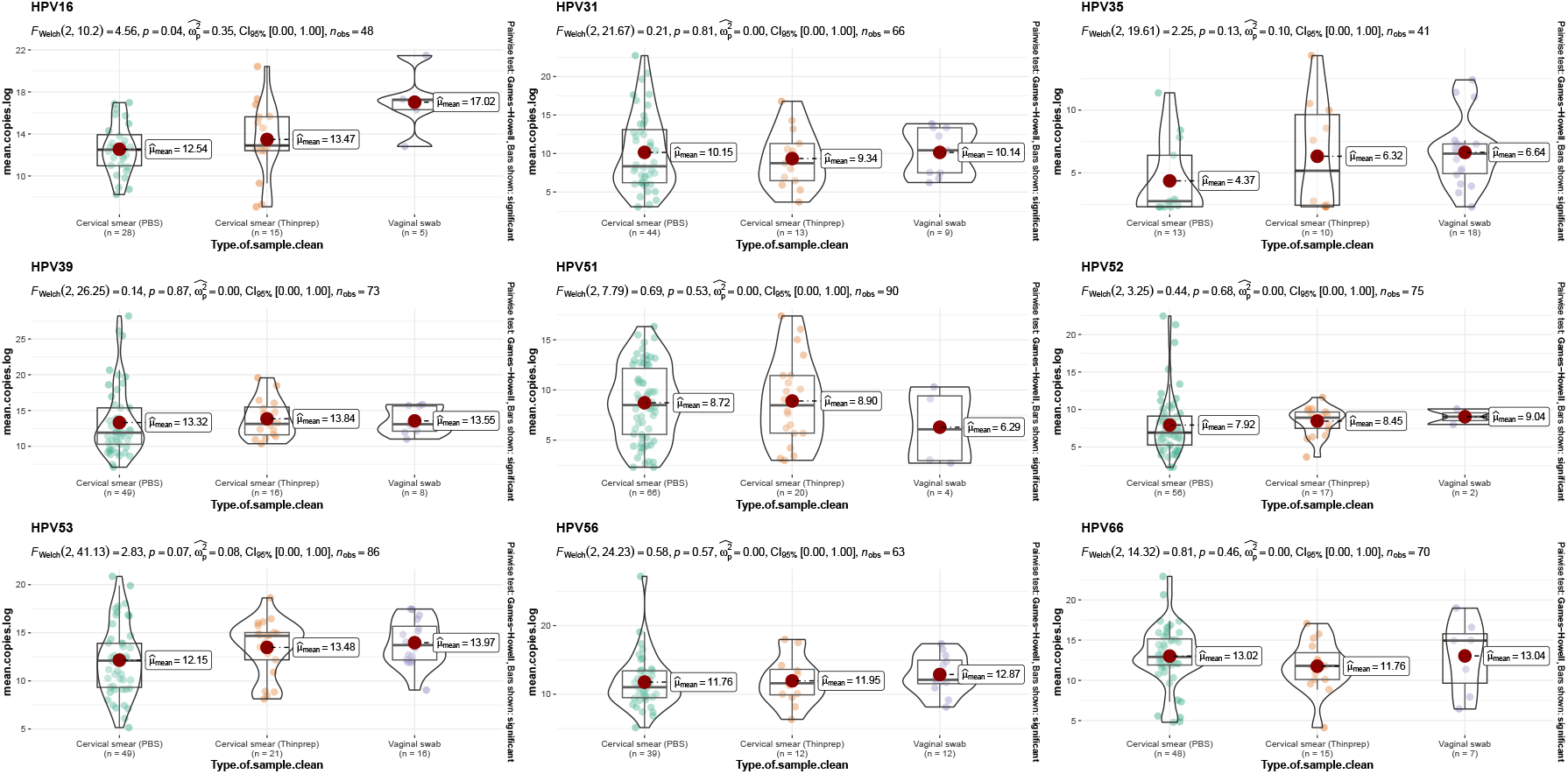
Viral load distribution per genotype and per type of sample. Violin plots of measured HPV viral load stratified by genotype (HPV18, HPV58, HPV59, and HPV68 are not shown). Red dots indicate the median viral load. For boxplots, the lower and upper hinges correspond to the first and third quartiles (the 25^th^ and 75^th^ percentiles). The violin plots display a mirrored density plot.

### Variance partitioning between the genotype, individual, and phylogenetic effects

Random effects allow our model to capture variations originating from three sources: the individual, the HPV genotype, and the evolutionary history, also referred to as phylogenetic signal. The latter takes into account the fact that some HPV genotypes are more genetically similar to others, which may lead to similar phenotypes. To determine the relative importance of these three sources, we calculated the variance partition coefficient of the posterior viral load. We found that the largest proportion of the variance originates from the genotype, with a median of 0.52, whereas both the individual level and the residuals showed a minor contribution to the variance partition, with respectively a median of 0.13 and 0.15. The effect of the phylogenetic signal was low. However, the distribution ranged from 0.1 to 0.9, with a peak near 0.1 and a median of 0.19, suggesting a heterogeneous distribution of the variation partition coefficient. Taken together, these results indicate that the majority of the variance for the posterior HPV viral load comes from the genotype level.

### Phylogenetic distance and HPV viral load

We further explored the correlation between genomic distance and virus load. For this, we represented a phylogenetic tree restricted to the 13 HPV genotypes in our dataset and coloured its leaves according to the median of the posterior viral load (Figure 5). The two genotypes with the highest virus load, namely HPV16 and HPV39, appear to be outliers among their clades. However, we also identify a clade composed of HPV56, HPV66, and HPV53 with similar posterior viral distribution. This feature probably explains the phylogenetic signal and heterogeneous distribution detected earlier (Figure 4).

**Figure 4.**
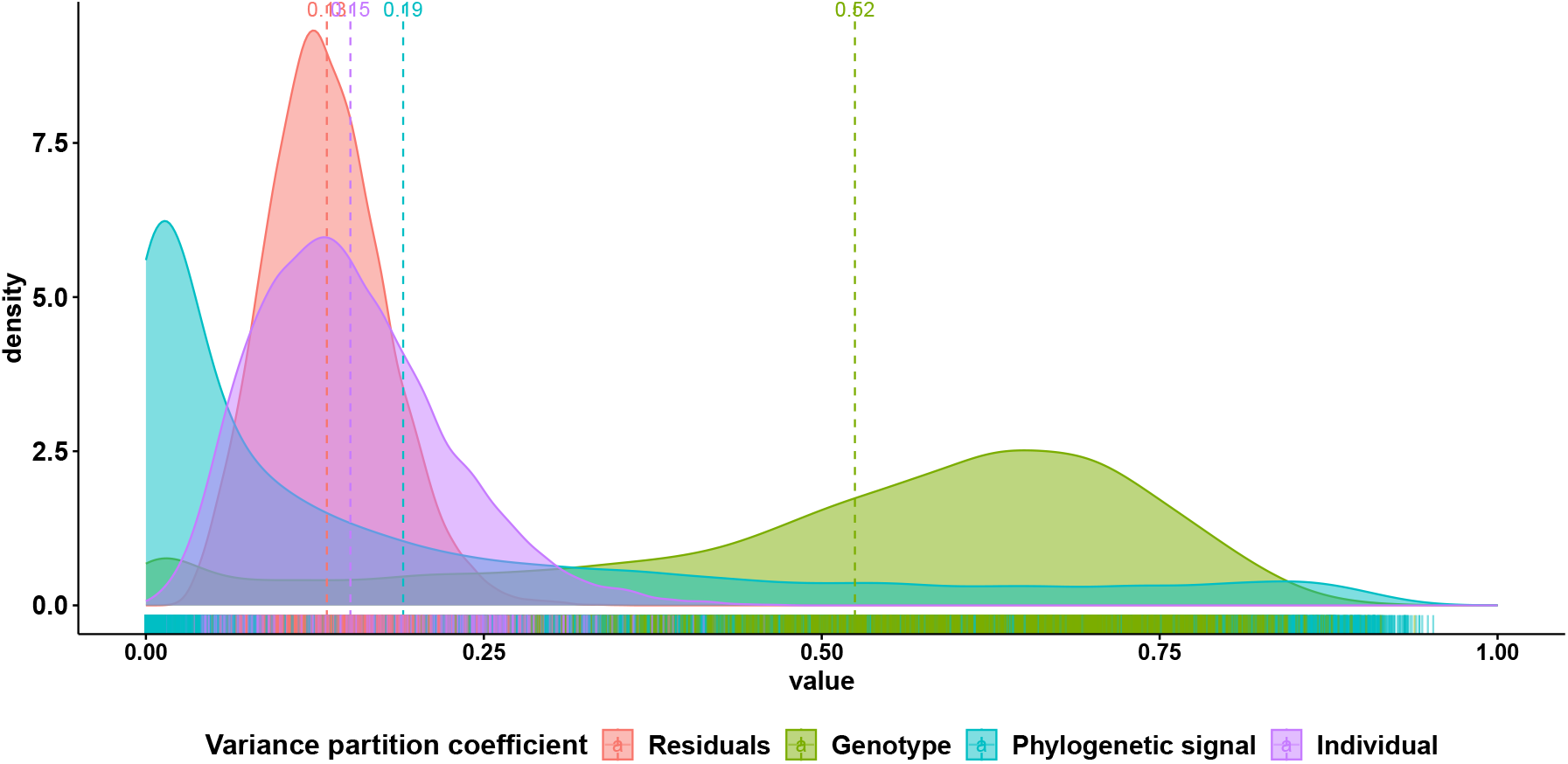
Distribution of the variance partition coefficients for the three random effects and the residuals of Model 1. Variance partition coefficients (VPC) were calculated for the individual, genotype, and phylogenetic signal random effects, as well as the residual variance (see the Methods for details). VPCs distributions are plotted with marginal rugs for each posterior draw. The median value for each distribution is indicated at the top of the color-matching dashed line.

**Figure 5.**
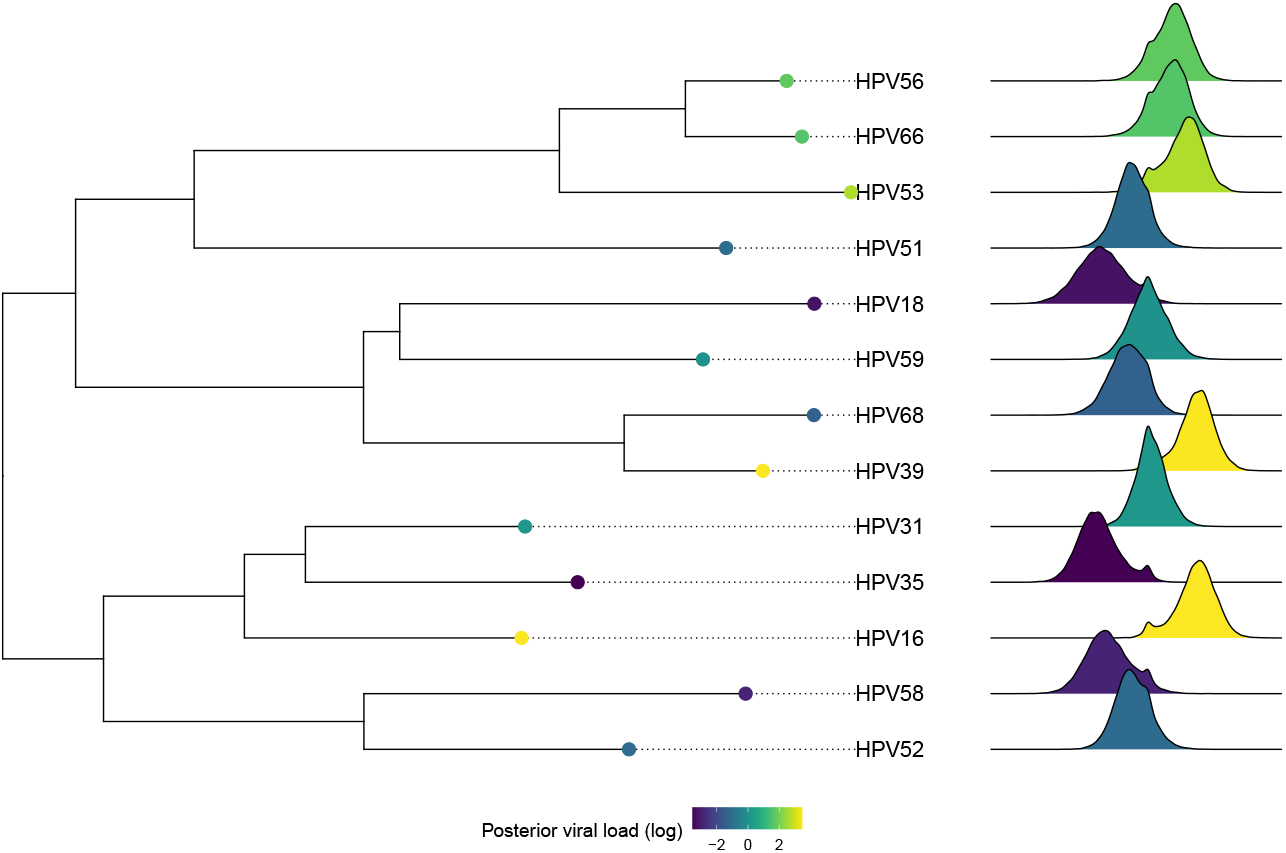
Phylogenetic distribution of HPV viral loads. Unrooted non-ultrametric HPV phylogenetic tree restricted to the 13 genotypes included in the study. The genotype-matching posterior viral load distributions are shown on the right. The tree leaves and the distributions are coloured according to the median posterior viral load estimated by Model 1.

## 3. Discussion

HPV viral load has been put forward as a biomarker for disease progression. However, the determinants of variation in HPV viral load are still poorly known. Here, we study the association between different covariates and the viral load by accounting for differences among individuals, HPV genotypes, and even genomic distance.

### Factors influencing per-cell viral load

Among the covariates included in our hierarchical Bayesian model as fixed effects, the strongest association with the virus load came from the type of sampling. We did not find an association between virus load and coinfection status, although both have been reported to the associated with increased persistence by earlier studies (oz et al. 2020). This is not necessarily inconsistent since virus load dynamics in non-persistent infections are poorly known and, for instance, viral competition for host resources or apparent competition via the immune system could generate such patterns (Murall et al. 2014). Nonetheless, it seems important to establish, especially for HPV16, for which the viral load is known to increase positively with disease progression.

### Sampling technique

Our results point toward a higher viral load in samples taken from vaginal swabs. Hence, a growing effort could be considered using this sampling method to identify its suitability for HPV screening. A case could be made about vaginal swabs being less invasive and so more easily acceptable than cervical smears for HPV screening. Given that in some European countries, HPV screening coverage is still low (source), and that uneasiness with the gynaecological examination has been cited as a common motive for missing the screening program, some have proposed that self-sampling using vaginal swabs could be an opportunity to increase screening program coverage. Our results show that vaginal swabs are not only efficient to measure HPV viral load but also seem to have a lower variability per-cell viral load. However, vaginal swabs do not sample the same region of the female reproductive tract as cervical smears, which may generate differences among HPV genotypes.

### Genotype effect on viral load

According to our model, the majority of the observed variance was explained by the HPV genotype. Strikingly, HPV16 was the genotype with the highest estimated viral load. Furthermore, the average differences in virus load between genotypes were high, with up to 4 orders of magnitude, which excludes potential differences in assay sensitivity. Overall, these results highlight the importance of considering the HPV genotype when trying to use viral load in diagnostics approaches or as a biomarker for disease progression. Another factor increasing per-cell viral load is the presence of a medium-risk HPV genotype. However, in our study, this category was represented by only two genotypes, namely HPV53 and HPV66. Given the discrepancy between other high-risk HPV genotypes, further results with other medium-risk HPV genotypes should be considered before asserting such a conclusion.

### Phylogenetic signal

One explanation for the differences between genotypes is that they originate from a shared ancestry and genomic similarity. Our results failed to identify any strong phylogenetic signal related to the estimated viral load. One exception was for a clade of three genotypes, HPV56, HPV66 and HPV53, which displayed a similar estimated viral load. One hypothesis that could explain this apparent outlier is the fact that studies have suggested that co-divergence of HPV16 sub-lineages with separate but closely related ancestral *Hominin* populations with subsequent host-switch events between archaic and modern human ancestral populations (Pimenoff et al. 2016).

An earlier study already identified a correlation between genomic distance and vaccine protection (Bogaards et al. 2019). One difference is that their phylogeny was inferred for the L1 gene because the virus-like particles from the vaccine are derived from it. We used the E1-E2 because it allows to group the oncogenic HPVs (Bravo and Alonso 2004).

### Perspectives

We showed that HPV virus load vary greatly among genotypes in our cohort, with HPV16 exhibiting the highest values. We also find that some techniques allow one to sample more viruses, an interesting feature being that this is particularly true for HPV16, the most oncogenic genotype by far. The association between virus load and oncogenicity is unclear. However, there are also strong variations within genotypes so finer genomic resolution would be needed. Furthermore, we only followed young women and, logically, did not detect any HSIL. Identifying associations between virus load and high-grade lesions would require cohorts of older women. We do show that such a study should pay particular attention to the HPV genotype and the sampling method. More generally, we show that swabs may be an interesting option for HPV16 detection, further supporting the idea to rely on self-sampling for cervical cancer screening.

One important question still not resolved with this study is whether HPV viral load could be used as a biomarker for disease screening. Given the technical challenges of the qPCR needed to deploy per genotype, it doesn’t seem realistic that it could be deployed as a first-step screening strategy. Nevertheless, on second intention, studies have suggested that for follow-ups, it could be valuable to know about the dynamic of the viral load. As such, it may help understand the dynamic of the infection, and could possibly reveal latent or clearing HPV infections. However, the present study suggests that vaginal swabs may be good candidates for specific HPV screening (especially HPV16), which raises the possibility of self-sampling strategies to monitor the progression of the viral load.

## 4. Materials and methods

### Clinical study description

The PAPCLEAR study took place in Montpellier (France) between 2016 and 2020 to study the natural history of HPV infections and further details about its protocol can be found in (Murall et al. 2019). Participants were between 18 and 25 years old at inclusion and had to report a new sexual partner within the last 12 months. The majority of them were university students. Further details about the cohort can be found in (Murall et al. 2020).

Here, we analyse samples collected at the inclusion visit (V1) and at the return visit (V2) four weeks later for participants who were infected by HPV genotypes for which we could quantify virus load using the quantitative Polymerase Chain Reaction (qPCR) protocol described below.

### Sample types and collection

All the samples were collected by a gynaecologist or midwife at the STI detection center (CeGIDD). Here, we analyse three types of samples: cervical smears collected using a cytobroom and put in PreservCyt medium (Hologic, Malborough, MA, USA), cervical smears collected using a cytobrush and put in PBS, and cervical swabs collected using eSwabs^TM^ (Copan, Murrieta, USA) and put in Amies medium.

### DNA extraction and quantification

For cytobroom samples in PreservCyt, we centrifuged 2mL (5min at 3 000g) and resuspended a pellet in 200µl of PBS. Elution was performed in 100µl of the kit buffer. For cytobrush samples in PBS, the solution was filtered and the filter was cleaned with RPMI. We then performed a centrifugation (10min at 514g). The pellet was resuspended in 200µL of PBS and a 20µL aliquot was used for DNA extraction. DNA was extracted using Qiamp DNA mini kit (Qiagen).

### qPCR

We implemented a uniplex version of the HPV genotype-specific method from Micalessi et al. (2012) and further described in Uysal et al. (2022).

### Vaginal microbiota composition

The microbiota metabarcoding was performed on 200 µL of vaginal swabs specimen stored at −80° in Amies medium. DNA extraction was performed using the MagAttract PowerMicrobiome DNA/RNA Kit (Qiagen). Next-generation sequencing of the V3-V4 region of the 16S gene Frank et al. (2008) was performed on an Illumina HiSeq 4000 platform (150 bp paired-end mode) at the Genomic Resource Center at the University of Maryland School of Medicine. Taxonomic assignment was performed using the internal software package SpeciateIT (https://github.com/Ravel-Laboratory/speciateIT) and the community state type (CST) was determined using the VALENCIA software package France et al. (2020).

### Evolutionary analyses

The HPV phylogeny was inferred using RAxML v.8.2.12 (Stamatakis 2014) from the concatenated E1 and E2 genes assuming a GTR substitution model.

### Statistical analyses

Statistical analyses were performed using R v.4.0.1.

The equations describing the Bayesian model are the following:

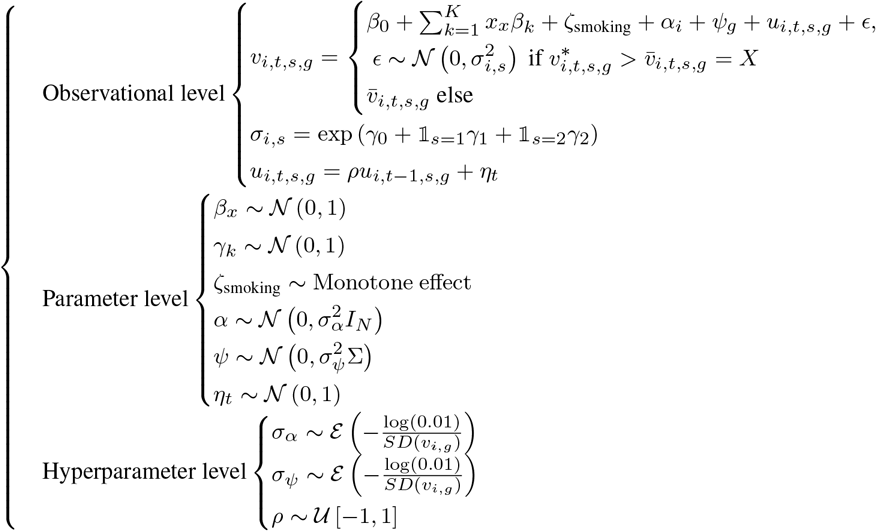

with *I*_*N*_ the identity matrix of size *N × N* and Σ a matrix encoding the phylogenetic distance.

### Ethics

The PAPCLEAR cohort study has been approved by the Comité de Protection des Personnes (CPP) Sud Méditerranée I (reference number 2016-A00712-49), the Comité Consultatif sur le Traitement de l’Information en matie`re de Recherche dans le domaine de la Santé (reference number 16.504), the Commission Nationale Informatique et Libertés (reference number MMS/ABD/ AR1612278, decision number DR-2016–488), the Agence Nationale de Sécurité du Médicament et des Produits de Santé (reference 20160072000007), and is registered at ClinicalTrials.gov under the ID NCT02946346.

## Data Availability

All data produced in the present study will be made available upon publication in a peer reviewed journal.

## Acknowledgements

This project was funded by the European Research Council (ERC) under the European Union’s Horizon 2020 research and innovation programme (grant agreement No 648963).

NT and OS were funded by the ANRS-MIE and TK was funded by the FRM.

Data used in this work were partly produced through the GenSeq technical facilities of the “Institut des Sciences de l’Evolution de Montpellier”. With the support of LabEx CeMEB, an ANR “Investissements d’avenir” program (ANR-10-LABX-04-01).

The authors acknowledge the ISO 9001-certified IRD i-Trop HPC (South Green Platform) at IRD Montpellier for providing HPC resources that have contributed to the research results reported within this paper. https://bioinfo.ird.fr/

## A Supplementary materials

**Figure S1.**
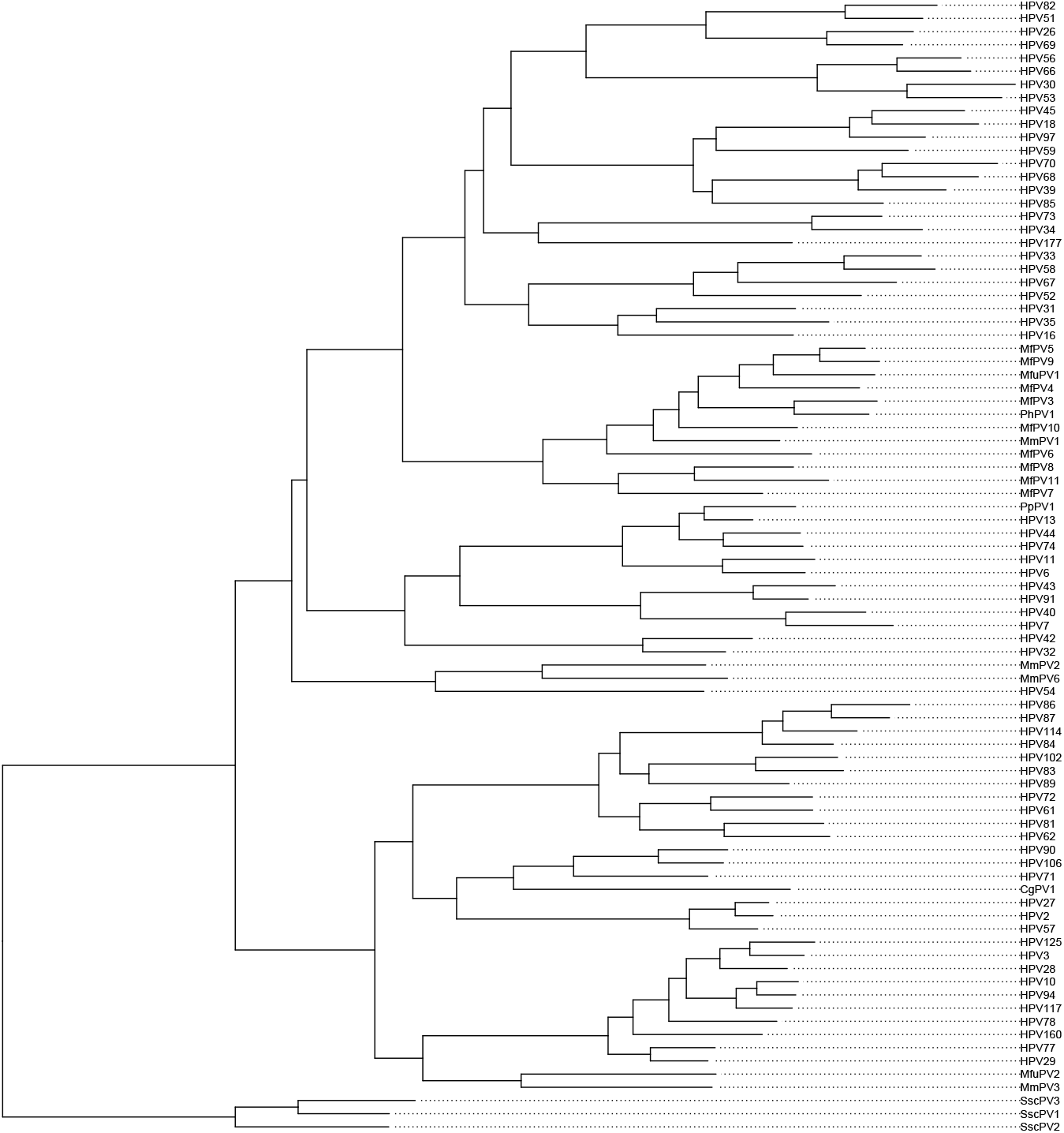
Phylogenetic tree of α-Papillomaviruses. Phylogenetic tree used to calculate the varicance::covariance matrix used in **??**

**Figure S2.**
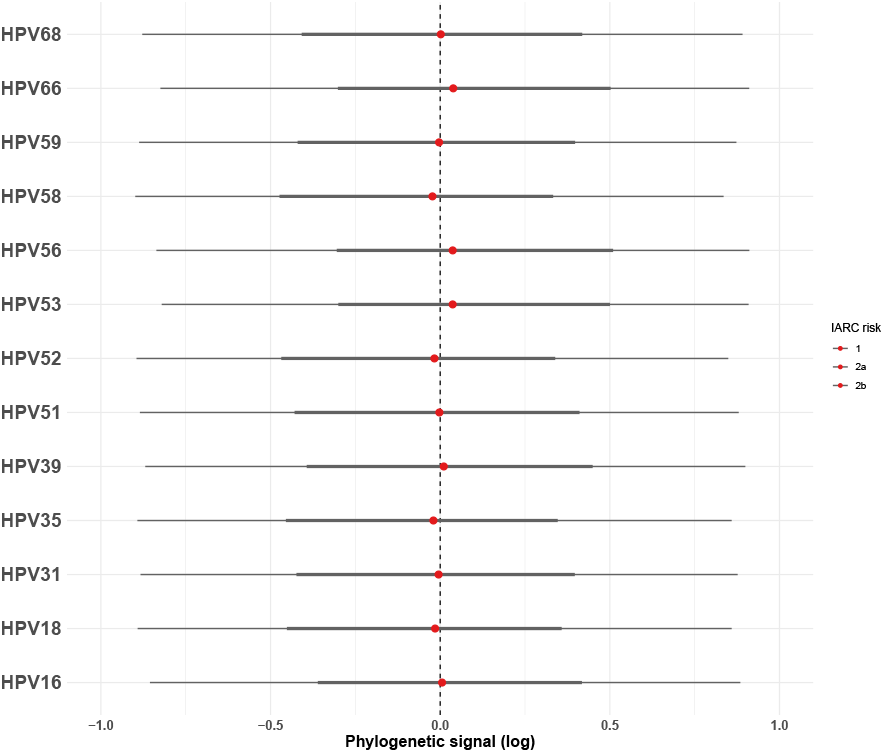
Estimated phylogenetic random effect for each genotype in the study. For each genotype included in the model (see 4.), posterior phylogenetic signal was estimated. The estimated 66% (thick line) and 95% (thin line) quantiles intervals are represented.

**Figure S3.**
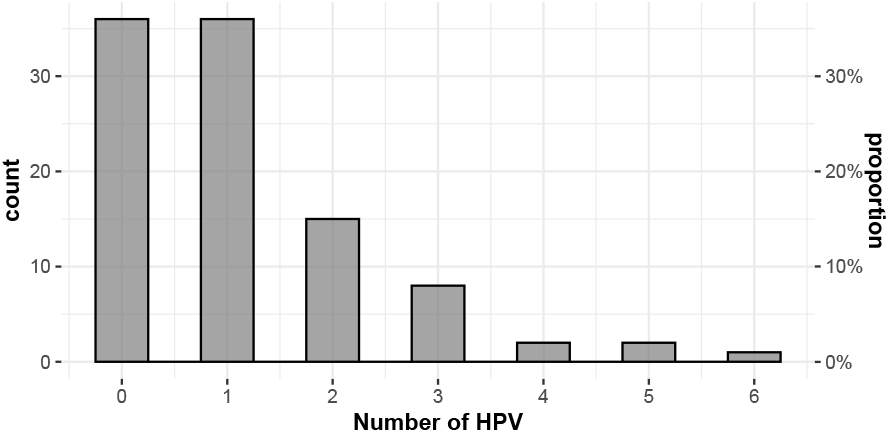
Distribution of HPV genotypes number per participant. Number of different HPV genotypes detected by LIPA

**Table S1.** Results for approximated LOO-CV. Loo-CV comparison of model [1] and model [2] which differ by the parameters included in the heteroscedasticity. Model [1] has heteroscedasticity on the type of sample parameters alone, whereas model [2] has heteroscedasticity for type of samples and genotype parameters.

| Heteroscedasticity | Parameter | Estimate | SE | Good | Ok | Bad | Very bad |
| --- | --- | --- | --- | --- | --- | --- | --- |
| 1 + Type of sample | ELPD | -1828.09422 | 24.111570 | 677 | 3 | NA | NA |
|  | Effective number of parameters | 66.02527 | 4.691630 | 677 | 3 | NA | NA |
|  | LOOIC | 3656.18843 | 48.223140 | 677 | 3 | NA | NA |
| 1 + Type of sample + genotype | ELPD | -1836.52944 | 26.330615 | 656 | 20 | 3 | 1 |
|  | Effective number of parameters | 91.03449 | 8.699127 | 656 | 20 | 3 | 1 |
|  | LOOIC | 3673.05888 | 52.661229 | 656 | 20 | 3 | 1 |

**Table S2.** Co-infections presents in the cohort study.

| Coinfections genotypes | Freq |
| --- | --- |
| HPV-51 - HPV-53 | 2 |
| HPV-53 - HPV-66 | 2 |
| HPV-16 - HPV-18 - HPV-51 | 1 |
| HPV-16 - HPV-31 - HPV-51 - HPV-52 - HPV-53 - HPV-66 | 1 |
| HPV-16 - HPV-31 - HPV-52 - HPV-56 | 1 |
| HPV-16 - HPV-39 - HPV-51 | 1 |
| HPV-16 - HPV-51 | 1 |
| HPV-16 - HPV-56 - HPV-66 | 1 |
| HPV-16 - HPV-59 | 1 |
| HPV-18 - HPV-51 - HPV-54 - HPV-68/73 | 1 |
| HPV-18 - HPV-53 - HPV-59 | 1 |
| HPV-18 - HPV-53 - HPV-66 | 1 |
| HPV-31 - HPV-35 | 1 |
| HPV-31 - HPV-45 - HPV-56 - HPV-66 - HPV-70 | 1 |
| HPV-31 - HPV-51 | 1 |
| HPV-31 - HPV-52 - HPV-53 - HPV-58 - HPV-68/73 | 1 |
| HPV-31 - HPV-58 | 1 |
| HPV-34 - HPV-66 | 1 |
| HPV-35 - HPV-43 | 1 |
| HPV-35 - HPV-51 - HPV-66 | 1 |
| HPV-35 - HPV-56 | 1 |
| HPV-39 - HPV-53 - HPV-68/73 | 1 |
| HPV-42 - HPV-66 | 1 |
| HPV-44 - HPV-52 - HPV-53 - HPV-39/68/73 | 1 |
| HPV-44 - HPV-52 - HPV-68/73 | 1 |
| HPV-44 - HPV-68/73 | 1 |
| HPV-45 - HPV-52 | 1 |
| HPV-51 - HPV-52 | 1 |
| HPV-51 - HPV-52 - HPV-53 | 1 |
| HPV-51 - HPV-52 - HPV-66 | 1 |
| HPV-51 - HPV-53 - HPV-56 | 1 |
| HPV-51 - HPV-56 | 1 |
| HPV-51 - HPV-56 - HPV-68/73 | 1 |
| HPV-52 - HPV-68/73 - HPV-74 | 1 |
| HPV-53 - HPV-54 | 1 |
| HPV-53 - HPV-56 | 1 |
| HPV-53 - HPV-59 | 1 |
| HPV-53 - HPV-70 | 1 |
| HPV-54 - HPV-59 | 1 |
| HPV-54 - HPV-68/73 | 1 |
| HPV-6 - HPV-16 - HPV-53 - HPV-39/68/73 | 1 |
| HPV-6 - HPV-31 - HPV-44 | 1 |
| HPV-66 - HPV-68/73 | 1 |

**Table S3.** HPV genotype presence in coinfections.

| HPV genotype presence in co-infections | Counts |
| --- | --- |
| HPV-51 | 24 |
| HPV-53 | 24 |
| HPV-66 | 18 |
| HPV-52 | 17 |
| HPV-68/73 | 15 |
| HPV-16 | 14 |
| HPV-54 | 11 |
| HPV-31 | 10 |
| HPV-39 | 10 |
| HPV-56 | 9 |
| HPV-35 | 8 |
| HPV-44 | 7 |
| HPV-18 | 6 |
| HPV-59 | 5 |
| HPV-43 | 4 |
| HPV-42 | 3 |
| HPV-45 | 3 |
| HPV-39/68/73 | 3 |
| HPV-6 | 2 |
| HPV-58 | 2 |
| HPV-70 | 2 |
| HPV-74 | 2 |
| HPV-34 | 1 |
| HPV-40 | 1 |

**Table S4.** Factors used in the best models selection by AIC.

| Variable | Status | Type | Levels |
| --- | --- | --- | --- |
| Viral load (per-cell normalised) | Response | Numeric |  |
| Sampling technique | Covariable | Factor | Cervical smears (Thinprep or PBS) or Vaginal swabs |
| HPV risk | Covariable | Binary | High or Medium |
| Co-infections | Covariable | Binary | Absent or Present |
| Gynaecologist | Covariable | Factor | A or B or C |

